# Scarlet Fever and Meteorological Exposures in Jiangsu, China: A Time-stratified Case-crossover Study

**DOI:** 10.1101/2025.03.11.25323804

**Authors:** Kai Wang, Wendong Liu, Hongfei Zhu, Yifan Tang, Hong Ji, Ying Wang, Liguo Zhu, Chengxiu Ling

## Abstract

**Background:** Increasing interest arises in the association between short-term meteorological exposure and scarlet fever risk, but the association with individual-level infection risk remains poorly understood.

**Methods:** We collected weather data from ERA5-Land (hourly, 9 km × 9 km) and aggregated into daily exposures to match all scarlet fever cases data in Jiangsu, reported the Nationwide Notifiable Infectious Diseases Reporting Information System from 2005 to 2023. We conducted a time-stratified case-crossover study with associations quantified by odds ratio (ORs) with confidence interval (CI) from conditional logistic regressions with distributed lag non-linear models.

**Results:** The odds ratios are generally significant with a 2–5 days lag, peaking at 3 days: 0.991 (95% CI: 0.986, 0.995) for temperature, 0.995 (95% CI: 0.994, 0.996) for relative humidity, 0.994 (95% CI: 0.990, 0.997) for total precipitation, 1.009 (95% CI: 1.005, 1.012) for solar radiation, and 1.088 (95% CI: 1.057, 1.120) for surface pressure. For non-linear effects, temperature showed a reversed U-shaped curve with peak risk between 15.17^°^C and 19^°^C and fluctuated risk at extremely low temperatures (below *−*5^°^C). Relative humidity posed a higher risk between 56% and 80%. Children aged over 6 exhibit greater susceptibility with stronger associations in temperature and surface pressure. Stronger associations were found in the post-COVID-19 era (2020–2023), particularly for temperature, solar radiation, and surface pressure.

**Conclusions:** Our study suggested significant non-linear associations between meteorological factors and scarlet fever risk, and provided some insights into the vulnerable children and the immune debt after COVID-19.

## 1 Introduction

Scarlet fever, caused by group-A *β*-hemolytic Streptococcus (Streptococcus pyogenes), is an acute respiratory infectious disease that predominately affects children aged 5 to 15 years [1]. Symptoms include sore throat, fever, headaches, swollen lymph nodes, angina, and a systemic red rash [2]. Historically, scarlet fever was prevalent in Europe and the United States during the 18th and 19th centuries, while its incidence declined in the 20th century [3]. However, in the past decade, the disease has resurgence in various regions in Asia and Europe, including Vietnam [3], South Korea [4], mainland China [1] and Hongkong City [5], Singapore [6], Poland [7] and England [8]. Unfortunately, there is no specific drug or vaccine available for scarlet fever, making surveillance and prediction of incidence important for public health.

Previous epidemiological studies found that scarlet fever incidence exhibits spatial clustering and seasonal fluctuations, often following a bimodal pattern [5, 9], as reported in Beijing City [10] and Jiangsu Province [11]. These findings imply the potentially important roles of meteorological effects in scarlet fever transmission. Ecological studies also identified meteorological factors such as sunshine duration, temperature, relative humidity, precipitation, and wind velocity as key contributors to the disease [12, 13]. However, these studies typically operate at the city or provincial level and often rely on generalized additive models (GAMs) that incorporate a set of explanatory variables and spatiotemporal effects. On the one hand, these studies often lack a fine-grained spatiotemporal analysis, which compromises the reliability of inferences and limits their generalizability to the individual level. On the other hand, while GAMs can capture complex relationships, they increase model complexity and computational burden, making it difficult to detect the effects of short-term exposure on acute events, and potentially leading to biases due to the omission of confounding factors.

Therefore, to explore meteorological effects while avoiding the aforementioned pitfalls, we apply the time-stratified case-crossover (TSCC) design [14, 15, 16] at the individual level and establish the corresponding conditional logistic regression models. Among various referent (control) selection strategies in case-crossover studies, the TSCC design, which matches case day with 3 to 4 control days based on the same day of the week within the same month of the same year, is recognized as the least biased method [17, 18]. This design effectively controls for long-term trends, seasonality, and individual-level confounders, simplifying thus the model while reducing the risk of confounding. In addition, beyond the conditional logistic regression [19] for individual infection, conditional Poisson (quasi-Poisson) regression [20, 21] for counts of cases is also widely applied in environmental epidemiology to estimate short-term associations between daily environmental exposures and acute health events, such as emergency department visits [22], perinatal mortality [23], hospital admission risks and costs [24], myocardial infarction mortality [25], chronic obstructive pulmonary disease (COPD) hospitalizations [26], hand, foot, and mouth disease (HFMD) [27], and 42 notifiable infectious diseases [28].

To the best of our knowledge, this is the first investigation of the effects of short-term meteorological exposures on individual scarlet fever infections. Our study is conducted in Jiangsu Province, China, with a comprehensive dataset of 36,912 cases among children aged 9 months to 79 years from 2005 to 2023. Our research provides a valuable opportunity to analyze the influence of various meteorological factors on the infection of scarlet fever. The findings will contribute to crucial epidemiological insights for the surveillance and prevention of scarlet fever and enhance the understanding of public health consequences of environmental conditions.

## 2 Methods

### 2.1 Data

#### 2.1.1 Study area and scarlet fever data

Jiangsu Province, consisting of 13 cities and 95 county-level divisions, is located at latitude 30^*°*^46^*′*^–35^*°*^08^*′*^N and longitude 116^*°*^21^*′*^–121^*°*^54^*′*^E, is an eastern-coastal province of China. It covers an area of 102,600 km^2^ with the Yangtze River passes through the southern part, borders the Yellow Sea in the east, Zhejiang Province and Shanghai in the south, Anhui Province in the west, and Shandong Province in the north. The majority of Jiangsu has a humid subtropical climate, while a humid continental climate appears in the north, and it undergoes a clear seasonal change with heavy and frequent rainfalls in spring and summer. Jiangsu is also the most densely populated province, with a population increased from 75 million to 80 million in the past decade.

As a class B notifiable infectious disease in China, cases of scarlet fever must be reported timely and directly to the Nationwide Notifiable Infectious Diseases Reporting Information System (NIDRIS), a system implemented formally in 2004 and covered all healthcare institutions throughout China [29]. The clinical manifestations of scarlet fever (ICD A38.01) are acute onset of fever, pharyngitis with strawberry tongue, red rash with a sandpaper feel, and itching. A throat swab culture and a skin smear stain are used to confirm Streptococcus (group A) bacteria infection. According to the NIDRIS, a total of 36,912 cases were reported in Jiangsu Province from January 2005 to December 2023. Each record of scarlet fever cases included gender, age, occupation, address, date of onset, and date of diagnosis.

#### 2.1.2 Meteorological data

The hourly data on air temperature, dew point temperature, solar radiation, eastward and northward wind components, surface pressure, and total precipitation are sourced from the ERA5-Land hourly dataset [30], with a spatial resolution of 9 km × 9 km. ERA5-Land is derived by replaying the land component of the European Centre for Medium-Range Weather Forecasts (ECMWF) ERA5 climate reanalysis, which combines model data with global observations into a complete and consistent dataset using physical laws.

The initial hourly data are aggregated into daily means. We also compute daily relative humidity using the Clausius–Clapeyron equation [31, 32], and daily wind speed is calculated as the square root of the sum of the squares of the eastward and northward wind components.

#### 2.1.3 Covariates

The transmission probability of scarlet fever can fluctuate due to short-term time-varying covariates. The transmission tends to be higher among students in school settings [13] and in neighbourhoods experiencing ongoing outbreaks. To account for these effects, we adjust the model using indicators for public holidays and summer/winter vacations, as well as incorporating the number of scarlet fever cases reported in the previous 7 days within each neighbourhood (the same 9 km × 9 km spatial grid for meteorological data).

#### 2.1.4 Study design and data process

We employ a TSCC design, where each individual serves as their own control [15, 16]. For each case, 3 to 4 control days are matched based on the same day of the week within the same month of the same year. This design facilitates the control for seasonality, long-term trends, and individual characteristics that are unlikely to change within a short time, such as demographic characteristics (e.g., sex, race, education, and weight) and living habits (e.g., smoking and drinking).

The residential addresses of the cases are matched to corresponding longitude and latitude coordinates, at least at the sub-district level. A total of 3546 (9.6%) cases are excluded due to mismatches (typos in records) or missing data in the sub-district information, leaving a final of 33,366 available for analysis.

Both the cases and controls are matched with meteorological variables (temperature, relative humidity, solar radiation, wind speed, surface pressure, and total precipitation) as well as covariates (holidays and the number of recent cases). Spatial alignment is achieved by assigning the grid values to the cases and controls, based on their geographical location within the grid. Temporal alignment is determined by matching the records according to the dates.

### 2.2 Statistical analysis

#### 2.2.1 Primary analysis

In the descriptive analyses, the distribution of scarlet fever cases is described based on the number of cases across different categories, including sex, age, season, and time periods (2005–2010, 2011–2019, 2020–2023). The three time periods are defined by two key events: the resurgence of scarlet fever in 2011 and the onset of COVID-19. The statistical level of all following analyses is set at 0.05 with two-tailed tests.

For the time-stratified case-crossover design, we use conditional logistic regression to assess the associations between meteorological conditions and the onset of scarlet fever. The model is adjusted for covariates, such as holidays and the number of recent cases (outbreak effect), and is implemented using the “survival” R package (R 4.2.2 version). The odds ratios (ORs) and their corresponding 95% confidence intervals (CIs) are used to quantify the associations in this study. ORs derived from the TSCC analysis can also be interpreted as relative risks (RRs), given that the selection of control periods is based on density sampling, making the exposure during control periods representative of the average exposure in the study population [33, 34].

For the lag effect, the maximum lag duration is set to a 10-day interval, which adequately captures the typical incubation period of scarlet fever (2 to 5 days). We consider both single-lag models (named single-lag1 to single-lag10), representing exposure conditions in 1 to 10 days prior to the day of interest, and moving average lag models (named ma-lag01 to ma-lag010), which capture the mean exposure levels over the 1 to 10 days preceding the day of interest.

To explore potential nonlinear associations, we incorporate a distributed lag nonlinear model (DLNM) framework that simultaneously accounts for non-linear exposure-response dependencies and delayed effects [35, 36, 37]. We compared basis matrices (natural cubic B-splines, B-splines, polynomials, linear thresholds, etc.) and necessary arguments (degrees of freedom, knots, degree, etc.) using the Akaike Information Criterion (AIC). A natural cubic spline with 3 degrees of freedom is chosen for the lag-response dimension, while a natural cubic spline with three internal knots placed at the 25th, 50th, and 75th percentiles of the location-specific meteorological variable distributions is employed for the exposure-response relationship within the cross-basis function. These analyses are performed within the “dlnm” R package (R 4.2.2 version).

To further assess the additive interactive effects of exposures on scarlet fever, we conduct conditional logistic models with the inclusion of each pair of exposures. This approach provides more insight than simply evaluating departures from multiplicativity when translating epidemiological findings into public health actions [25, 38]. The interactions are evaluated using relative excess odds due to interaction (REOI), attributable proportion due to interaction (AP), and the synergy index (S), with the following formulas:

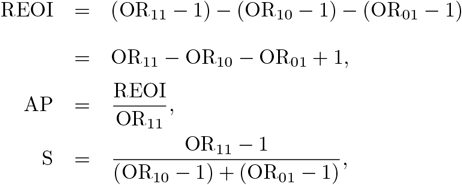

where OR_11_ refers to the simultaneous exposures, derived as the product of ORs of marginal and interaction effects in the model. The terms OR_01_ and OR_10_ refer to the single exposures, which are called as the marginal ORs.

The cases REOI = 0, AP = 0 and S = 1 indicate no interaction between the meteorological exposures. Positive values of REOI and AP, and S *>* 1 suggest synergistic interactions, meaning the combined effect exceeds the sum of individual effects. Conversely, negative values for REOI and AP, and S *<* 1 suggest antagonistic interactions, where the combined effect is weaker than the sum of the individual effects. Note that these metrics are valid only for cases where ORs *>* 1 for both exposures. For exposures with ORs *<* 1, we assign a negative sign to the exposure (to convert the ORs to *>* 1), which is interpreted as the change in ORs due to the decrease in exposures.

#### 2.2.2 Subgroup analysis

Considering the potential differences in meteorological effects across subgroups, we conduct subgroup analyses based on sex (male and female), age groups (0–6, 6+), and time periods (before and after the epidemic surge in 2011, and before and after COVID-19). These analyses are performed using the same conditional logistic regression model and DLNM framework.

#### 2.2.3 Sensitivity analysis

To assess the robustness of the results, we conduct several sensitivity analyses for DLNM: (a) change the degrees of freedom (to 4) for the natural splines in the lag responses. (b) change places of internal knots at the 33rd and 66th percentiles for meteorological exposures. (c) change the maximum lag period from 10 to 7 days.

## 3 Results

Table S1 summarizes the characteristics of scarlet fever patients across the groups. Of the total number of 33,366 scarlet fever cases, the majority of the patients are aged between 3 and 9 (84.6%), more than half are males (61.0%), and 67.4% occurred between the resurgence in 2011 and the onset of COVID-19, with an average annual incidence of 2499.8 cases. Seasonal peaks are observed in spring (32.8%) and winter (28.0%). Figure 1 shows the spatial distribution of all scarlet fever cases and the monthly incidence from January 2005 to December 2023. A higher spatial concentration of cases is observed in southern Jiangsu Province, with a significant increase in temporal incidence rates since the resurgence in 2011, reaching peaks in 2019.

**Figure 1:**
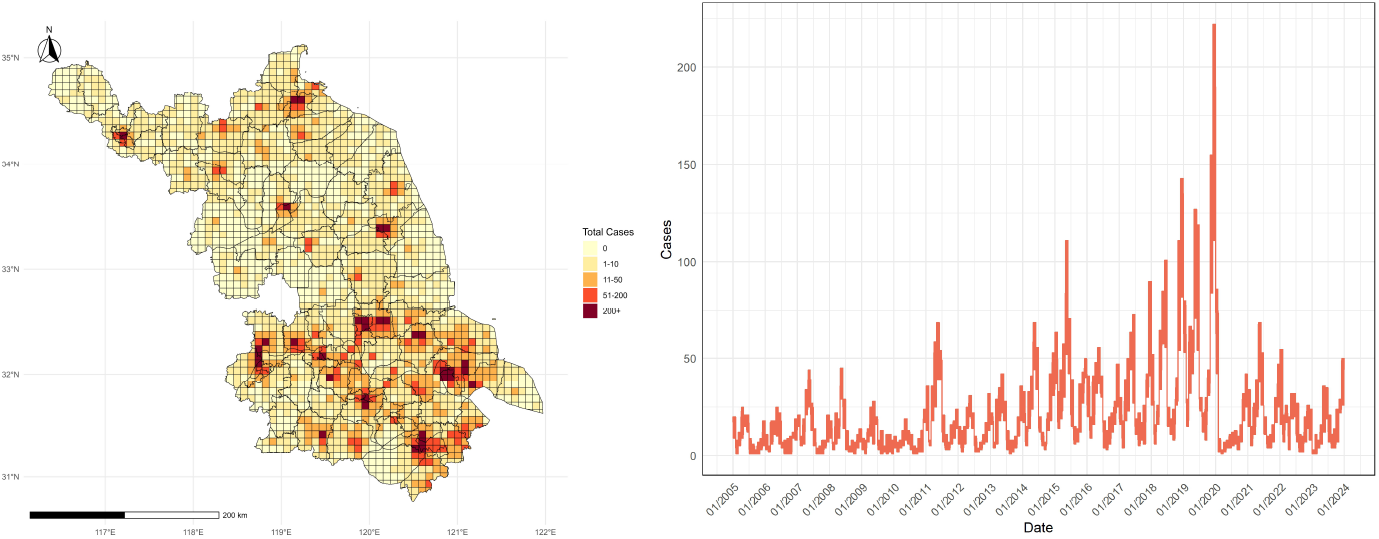
Spatial distribution (left) of the total number of cases in grids (9 km× 9 km) and temporal trend (right) of monthly incidence.

Figures 2 and S1 present the spatial distribution of mean levels and time-series plots of variations in meteorological exposures. Spatially, temperature, relative humidity, and total precipitation generally increase from north to south, and surface pressure decreases from east to west, while mean wind speed and solar radiation are higher along the eastern coast, as well as around Hongze and Taihu Lakes, which are prominent hot spots in the west and south. The temporal trends remain relatively stable, with consistent seasonal fluctuations throughout the study period (Figure S1).

**Figure 2:**
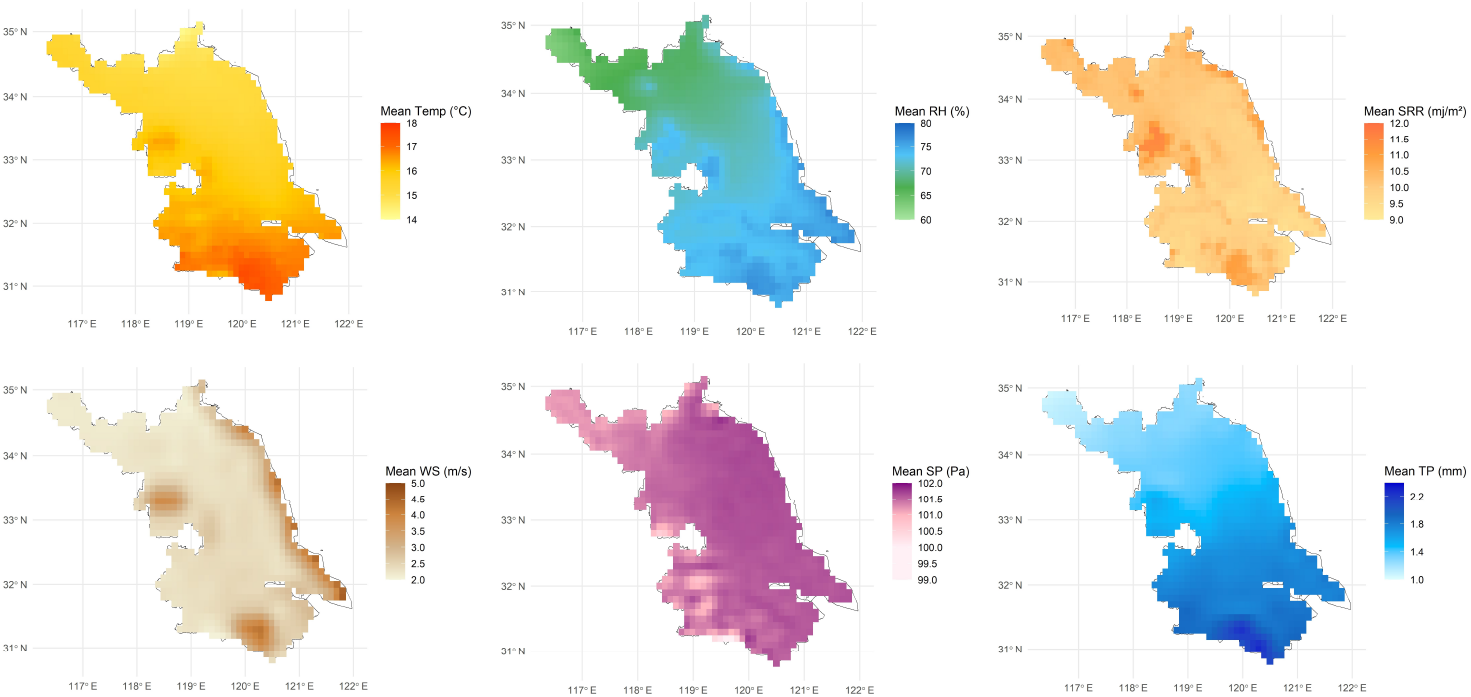
Spatial plots of means of all meteorological exposures in Jiangsu Province, China. Reference of mean levels (mean, standard deviation) are temperature (15.88, 9.35), relative humidity (71.57, 13.16), solar radiation (10.06, 4.45), wind speed (2.54, 1.23), surface pressure (101.48, 0.98), and total precipitation (1.58, 4.39).

The results of conditional logistic regression for the association between single exposures and scarlet fever are presented in Figure 3. The adjusted holiday and outbreak effects are all statistically significant across all models, with roughly equivalent OR estimates of 0.92 (95% CI: 0.87, 0.97) and 2.15 (95% CI: 2.12, 2.18), respectively. In single-lag models, significant negative associations were observed between exposures and scarlet fever from lag2 to lag6 for temperature, lag0 to lag5 for relative humidity, and lag0, lag2, and lag3 for total precipitation. Positive associations were found from lag0 to lag3 for solar radiation and from lag1 to lag6 for surface pressure. Similar results of exposures were also indicated by the ma-lag models, where the relative humidity, solar radiation, and surface pressure become consistently significant for all lags (lag01 to lag010).

**Figure 3:**
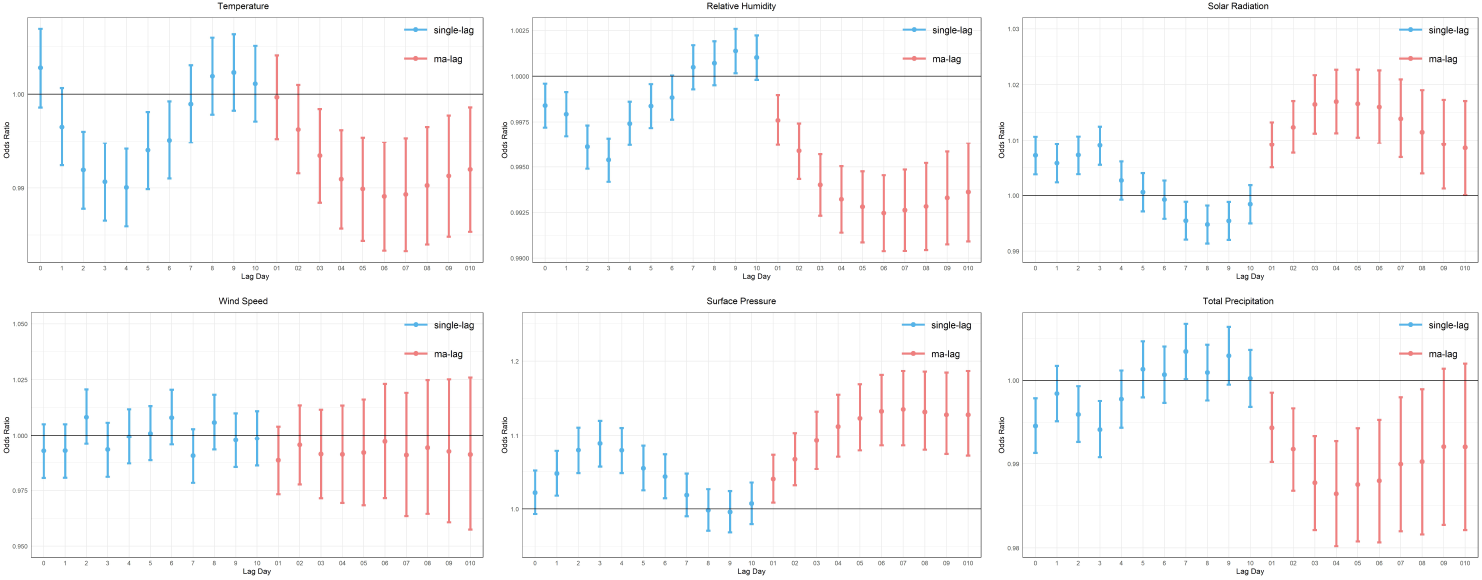
Associations indicated by ORs between six meteorological factors and scarlet fever in single-lag and ma-lag models.

The three-dimensional plots show the entire OR surface between the exposures and case at single-lag days (Figure S2) and ma-lag days (Figure S3). The estimated effects are non-linear for all the exposures, where only similar patterns in temperature were found in the two different lag models.

In the subsequent analysis, we primarily focus on the single-lag3 and ma-lag04 models, which align with the incubation period of scarlet fever (2 to 5 days). For the lag-3 model, the estimated ORs for a one-unit increase in each of the six exposures are as follows: 0.991 (95% CI: 0.986, 0.995), 0.995 (95% CI: 0.994, 0.996), 1.009 (95% CI: 1.005, 1.012), 0.993 (95% CI: 0.981, 1.005), 1.088 (95% CI: 1.057, 1.120), and 0.994 (95% CI: 0.990, 0.997). For the ma-lag04 model, the estimated ORs are 0.991 (95% CI: 0.986, 0.996), 0.993 (95% CI: 0.991, 0.995), 1.017 (95% CI: 1.011, 1.022), 0.991 (95% CI: 0.969, 1.014), 1.112 (95% CI: 1.071, 1.155), and 0.986 (95% CI: 0.980, 0.993).

Figures 4 and S4 display the exposure-response relationships for each meteorological factor across both lag models, with reference levels centred at median values. In the single-lag3 model, temperature exhibits a reversed U-shaped curve, where OR values exceed 1 between 15.17 (median) and 19 but fall below 1 outside this range. Relative humidity demonstrates ORs below 1 for values under 48. Solar radiation and total precipitation similarly exhibit ORs below 1 for values exceeding 17.5 and 15.1, respectively. Wind speed has elevated ORs at both low and high levels, while surface pressure generally decreases in OR with minor fluctuations as values increase.

**Figure 4:**
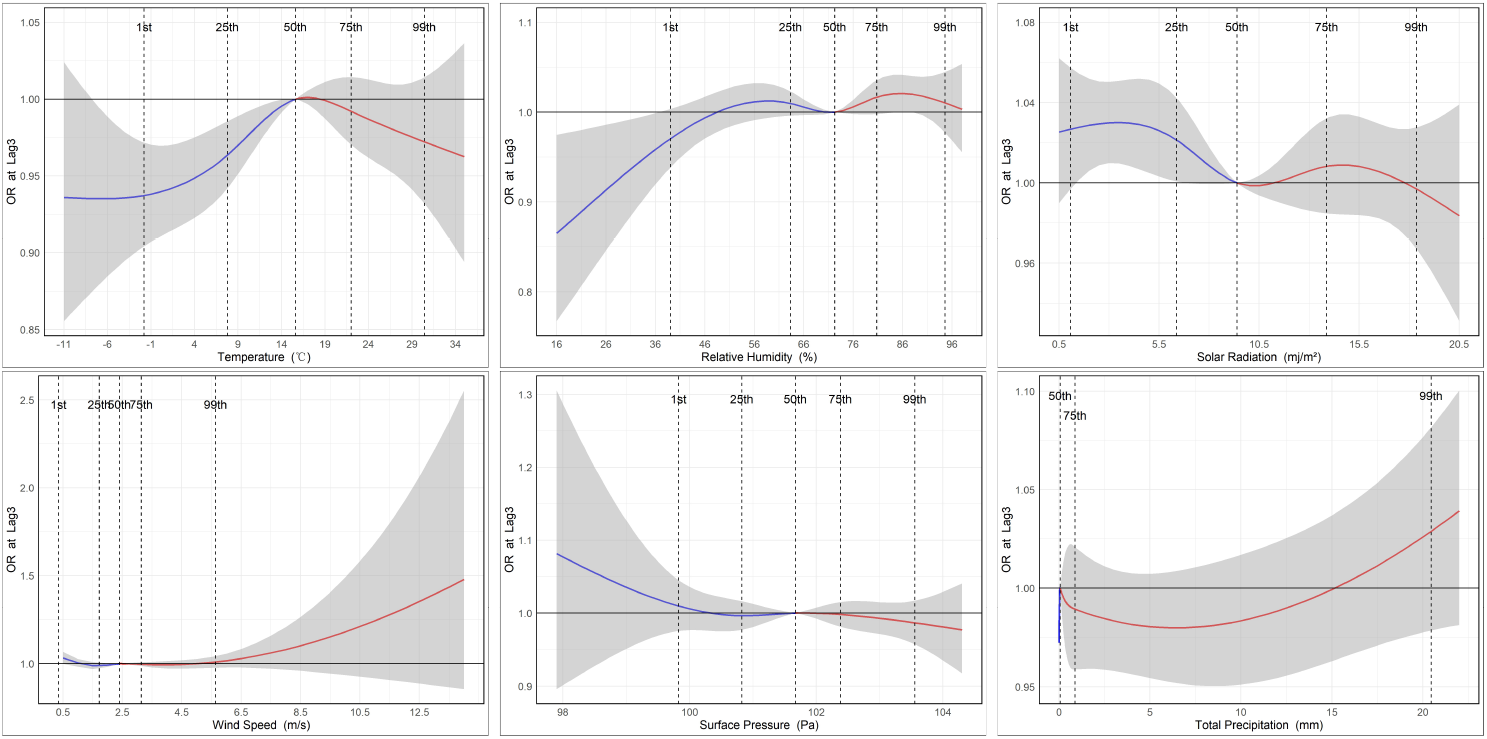
Exposure curves for non-linear relationship in the DLNM single-lag3 model.

In the ma-lag04 model, the temperature curve maintains its pattern, except at extremely low temperatures (below −5), where ORs rise above 1, although this effect is not significant. Relative humidity shows ORs below 1 for values under 46 and between 72 and 91. Solar radiation and surface pressure both present ORs below 1 at moderate values (7.7 to 9.6 for solar radiation and 98.9 to 101.7 for surface pressure). Wind speed and total precipitation exhibit relatively stable trends, remaining close to OR = 1 throughout.

Tables S1 and S2 summarize the interaction results for the 15 pairs of meteorological exposures in the single-lag3 and ma-lag04 models, along with the calculated REOI, AP, and S values with 95% CIs. Among the pairs, significant synergistic interactions (S *>* 1) are found between temperature and all other exposures, wind speed and humidity (surface pressure), as well as surface pressure and total precipitation. Antagonistic interactions (S *<* 1) are observed for wind speed and solar radiation. Some of these interactions are further supported by significant REOIs and APs, including the pairs involving temperature and humidity (wind speed, surface pressure), as well as wind speed and surface pressure.

Table 1 presents the subgroup analysis results from the single-lag3 model, with comparable findings indicated by the ma-lag04 model in Table S3. Children aged over 6 (13,409 samples) exhibit greater susceptibility to meteorological exposures compared to those under 6 (19,957 samples), with stronger associations observed for temperature (OR = 0.988, 95% CI: 0.983–0.994) and surface pressure (OR = 1.117, 95% CI: 1.077–1.159), while the other effects are similar. Gender-based analysis reveals consistent patterns for males and females, although the smaller female sample size (13,003 samples) may explain the discrepancy in precipitation significance. Regarding time periods, stronger associations are found in the post-COVID-19 era (2020–2023), particularly for temperature (OR = 0.989, 95% CI: 0.980–0.999), solar radiation (OR = 1.014, 95% CI: 1.005–1.023), and surface pressure (OR = 1.134, 95% CI: 1.060–1.212). In contrast, the pre-resurgence period (2005-–2010) shows fewer significant associations, suggesting evolving meteorological impacts on scarlet fever risk over time.

**Table 1:** Subgroup analysis of age group, gender, and periods, based on single-lag 3 model.

| Groups |  | Temperature | Relative humidity | Solar radiation | Wind speed | Surface pressure | Total precipitation |
| --- | --- | --- | --- | --- | --- | --- | --- |
|  |  | OR (95% CI) | OR (95% CI) | OR (95% CI) | OR (95% CI) | OR (95% CI) | OR (95% CI) |
| Age | 0–6 | 0.994 (0.987, 1.001) | 0.996 (0.994, 0.998) | 1.011 (1.005, 1.016) | 0.997 (0.977, 1.017) | 1.046 (0.999, 1.096) | 0.994 (0.988, 0.999) |
|  | 6+ | 0.988 (0.983, 0.994) | 0.995 (0.994, 0.997) | 1.008 (1.003, 1.012) | 0.991 (0.975, 1.007) | 1.117 (1.077, 1.159) | 0.995 (0.990, 0.999) |
| Gender | Male | 0.989 (0.984, 0.995) | 0.996 (0.995, 0.998) | 1.007 (1.002, 1.011) | 1.001 (0.985, 1.017) | 1.095 (1.055, 1.136) | 0.993 (0.989, 0.997) |
|  | Female | 0.993 (0.986, 0.999) | 0.994 (0.992, 0.996) | 1.013 (1.007, 1.018) | 0.982 (0.962, 1.001) | 1.077 (1.029, 1.128) | 0.996 (0.991, 1.001) |
| Time period | 2005–2010 | 0.989 (0.978, 1.001) | 0.992 (0.989, 0.995) | 1.008 (0.999, 1.018) | 1.001 (0.968, 1.035) | 1.084 (1.004, 1.170) | 0.991 (0.981, 1.001) |
|  | 2011–2019 | 0.991 (0.986, 0.996) | 0.997 (0.995, 0.998) | 1.008 (1.004, 1.012) | 0.989 (0.974, 1.004) | 1.077 (1.040, 1.116) | 0.995 (0.991, 0.999) |
|  | 2020–2023 | 0.989 (0.980, 0.999) | 0.993 (0.990, 0.996) | 1.014 (1.005, 1.023) | 0.997 (0.967, 1.027) | 1.134 (1.060, 1.212) | 0.994 (0.986, 1.002) |

Sensitivity analysis validated the robustness of our main results. Results of DLNM (Figures S5 ∼ S10) were robust for employing alternative degrees of freedom for the lag responses, places of internal knots for meteorological exposures, and different maximum lag days.

## 4 Discussion

Meteorological factors such as temperature, humidity, wind speed, and precipitation have been shown to significantly influence the occurrences and transmissions of both scarlet fever [12, 13] and other infectious diseases, such as streptococcal pharyngitis [39], tuberculosis [40], and HFMD [27, 41]. Recent ecological research has increasingly focused on exploring non-linear relationships using spline methods [42], and also accounting for delayed effects through DLNMs [13, 43], with broad applications in studies of diseases such as malaria [44] and dengue fever [45].

To the best of our knowledge, this is the first individual-level epidemiological study employing a case-crossover design to examine the association between meteorological exposures and scarlet fever risk. Our study includes all recorded scarlet fever cases in Jiangsu province from 2005 to 2023, encompassing periods of resurgence and the COVID-19 pandemic.

In this analysis, we found that generally lower temperatures are associated with an increased risk of scarlet fever infection, consistent with findings from Hong Kong [12]. The exposure-response analysis revealed a non-linear relationship with temperature: temperatures above 20 ^°^C and below 12 ^°^C are linked to reduced infection risk, while extremely low temperatures (below −5 ^°^C) exhibit fluctuating effects. These patterns align with findings from Guangzhou City in southern China [13], although the overall lower magnitude observed in our study can be attributed to the northern location of Jiangsu province. The reduced risk at higher temperatures, on the one hand, may result from reductions in outdoor bacterial levels, where protein denaturation and bacterial inactivation inhibit propagation [46, 47]. On the other hand, it can also be linked to human behaviour, as hot weather discourages outdoor activities, thereby reducing person-to-person transmission. At extremely low temperatures, increasing transmission risks could arise from both decreased human immune function and increased indoor gatherings, particularly during winter festivals. Conversely, decreased bacterial activity due to reduced cell membrane fluidity at low temperatures may mitigate risks [13]. These findings highlight the need for further investigation into the interplay of cold stress, immune resistance, and bacterial viability in shaping infection trends.

Lower relative humidity and total precipitation levels were associated with an increased risk of scarlet fever, consistent with observations across mainland China [9]. Relative humidity, in particular, may affect the ability of streptococcus pyogenes to produce toxins and enzymes, as well as influence transmission dynamics. High humidity can cause small respiratory droplets to absorb water, reducing the time pathogens remain airborne, which aligns with an inverse association between humidity and scarlet fever risk. The exposure-response relationship with relative humidity showed a clear nonlinear pattern in our study, with relative risk increasing as the humidity rose to approximately 60%, then declining slightly at higher levels. This pattern is consistent with the findings in Guangzhou [13] and supported by biological evidence. Low relative humidity can inhibit microbial activity, suppressing physiological processes such as metabolism in bioaerosols [47]. Meanwhile, higher humidity prevents dust from rising from wet surfaces, limiting bacterial introduction into the atmosphere.

Precipitation, as explored in previous studies, may influence scarlet fever incidence indirectly, primarily mediated by changes in human behaviour and reduce airborne bacterial loads by removing bacteria that adhere to particles through relative humidity [48]. The exposure-response curves also reflect this less significant relationship between total precipitation and scarlet fever risk. This discrepancy highlights the need to explore further how meteorological factors interact with environmental and behavioural mediators to influence scarlet fever transmission.

We found positive associations between solar radiation and surface pressure with scarlet fever incidence in Jiangsu, consistent with the findings in Hong Kong, Beijing, Hefei, and across 31 administrative regions [9, 12, 49]. For solar radiation, the positive effect decreases with fluctuation as levels rise, eventually turning negative at very high levels. This aligns with the weak positive association discussed in [9], where prolonged sunshine hours and high ultraviolet radiation were likely to reduce bacterial loads by damaging bacterial DNA. Surface pressure is closely linked to cold airflow, which enhances air diffusion and may facilitate the release and transport of bacteria, potentially increasing bacterial abundance in the atmosphere [13]. Our exposure-response curves further highlight a potentially stronger association at extremely low surface pressure levels. This may be attributed to the fact that low pressure can reduce the partial pressure of oxygen in the body, impairing physiological functions and weakening immune defences, thereby increasing susceptibility to infections [50].

The subgroup analysis revealed that children aged over 6 are more susceptible to meteorological conditions. This heightened susceptibility may be attributed to behavioural and social factors, as children in this age group are often more active outdoors, engage in more group activities, and attend school regularly, where close contact facilitates disease transmission. Stronger effects from meteorological exposures were also observed in the post-COVID-19 era, likely due to weakened immune systems and other adverse health outcomes following COVID-19 infections [51]. This phenomenon is linked to the concept of “immunity debt”, where the lack of immune stimulation—resulting from reduced circulation of microbial agents and lower vaccine uptake, particularly during COVID-19-related nonpharmaceutical interventions—may lead to an increased vulnerability to future outbreaks [52, 53].

Additionally, the general meteorological effects become significant with a lag of 3 to 5 days. This delay may be explained by the time required for the growth and reproduction of streptococcus pyogenes, as well as the incubation period from infection to the onset of scarlet fever symptoms [9, 54]. This temporal lag highlights the complex dynamics between environmental conditions and the biological processes underlying infection and disease progression. Understanding these delayed effects is crucial for anticipating outbreaks and implementing timely public health interventions.

As an individual-level study with a TSCC design, this study allows us to control the variations in socioeconomic status, health services, and hygiene, offering more credible and precise insights into how environmental conditions influence infection. Our approach also focuses on daily meteorological variables, enabling a more detailed investigation of acute effects. By examining short-term exposure-response relationships, we capture the immediate impact of meteorological factors on disease risk, which partially supports the findings from previous ecological studies [9, 12, 13, 49]. More importantly, as an epidemic case, we examine the meteorological influence by adjusting for the compound effect of contagious diseases, specifically the outbreak effect, in our analysis. To our knowledge, this adjustment is the first time incorporated into TSCC studies. The consistently significant outbreak effects across all models validate our approach, suggesting that neglecting this factor could lead to biases in estimating the meteorological effects, particularly in the context of infectious diseases with transmission dynamics.

However, several limitations of this study need to be addressed. First, approximately 10% of the records were excluded due to mistakes in address information, which is also a common issue in similar studies. Second, while we accounted for potential time-varying covariates, others were not included. For instance, multiple air pollutants and their interactions with meteorological conditions were not considered, while they were shown to influence scarlet fever incidence [55]. Third, our analysis relied on daily mean exposures, but other metrics such as daily maxima, minima, or ranges may also play an important role and require further investigation. Lastly, the absence of a standardized diagnostic criterion for scarlet fever across different regions could contribute to under-reporting or over-reporting of cases. These limitations highlight the need for future studies to address these gaps for a more comprehensive understanding of the factors influencing scarlet fever transmission.

## 5 Conclusion

Our study used a case-crossover design and identified significant associations between meteorological factors and scarlet fever risk, with non-linear exposure-response patterns. Subgroup analyses revealed greater susceptibility in older children and stronger effects post-COVID-19, reflecting potential behavioural and immunity-related influences. By addressing the limitations of previous ecological studies, this research provides evidence for incorporating meteorological and demographic factors into scarlet fever prevention strategies.

## Supporting information

All supplemental materials

## Data Availability

All data produced in the present study are available upon reasonable request to the authors.

## Declarations

### Ethics approval and consent to participate

The study is approved by the Xi’an Jiaotong-Liverpool University Research Ethics Committee (0010000089620241213145342).

### Availability of data and materials

The data that support the findings of this study are available from the Jiangsu Provincial Center for Disease Control and Prevention, but restrictions apply to the availability of these data, which were used under license for the current study and so are not publicly available. Data are, however, available from the authors upon reasonable request and with permission of the Jiangsu Provincial Center for Disease Control and Prevention.

### Competing interests

The authors declare that they have no competing interests.

### Funding

This research was supported by the Jiangsu Province 333 project, National Key R&D Program of China (2024YFC2310403). This research was partially supported by the Top Talent Awards Project Fund (RDF-TP-0023, RDF-TP-0030) and Post-graduate Research Fund (PGRS2112022) at Xi’an Jiaotong-Liverpool University.

### Authors’ contributions

Chengxiu Ling, Liguo Zhu, and Ying Wang contributed to the research design, conceptualization and validation; Kai Wang contributed to writing (original draft), formal analysis and visualization; Wendong Liu contributed to writing and methodology review; Hongfei Zhu contributed to data process, data resource and validation; Yifan Tang and Hong Ji contributed to methodology, visualization, and review. All authors participated in the revision and approved the manuscript.

## Acknowledgement

We thank the editors and anonymous reviewers for their helpful remarks.

## Consent to participate

Not applicable.

### Consent to publish

Not applicable.

## Notes

### Competing Interest Statement

The authors have declared no competing interest.

### Author Declarations

Ethics committee of Xi'an Jiaotong-Liverpool University Research Ethics Committee gave ethical approval for this work (0010000089620241213145342).

