## Supplementary material for "Scarlet Fever and Meteorological Exposures in Jiangsu, China: A Time-stratified Case-crossover Study": All supplemental materials

#### 1 Figures

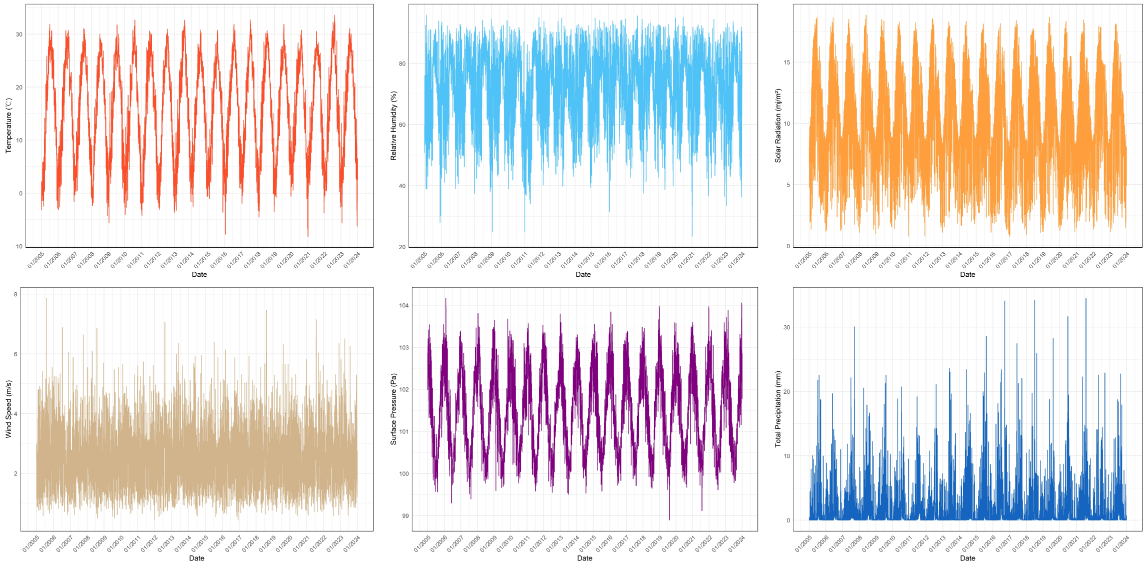


Figure S1: Temporal trend of meteorological exposures

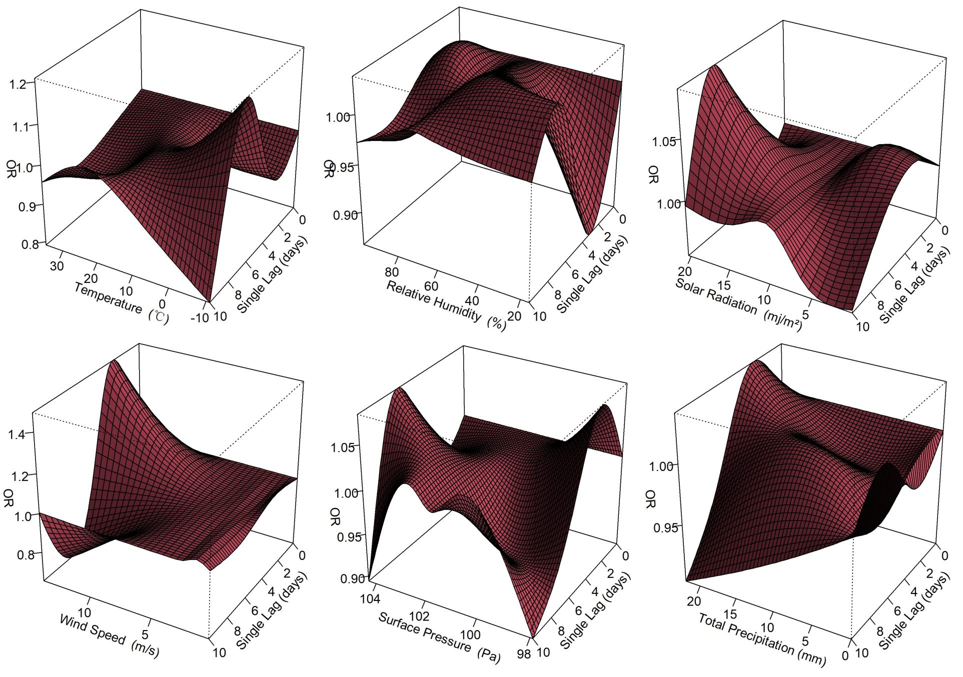


Figure S2: Three-dimensional plots for OR surface with single-lag models and non-linear effect in DLNM.

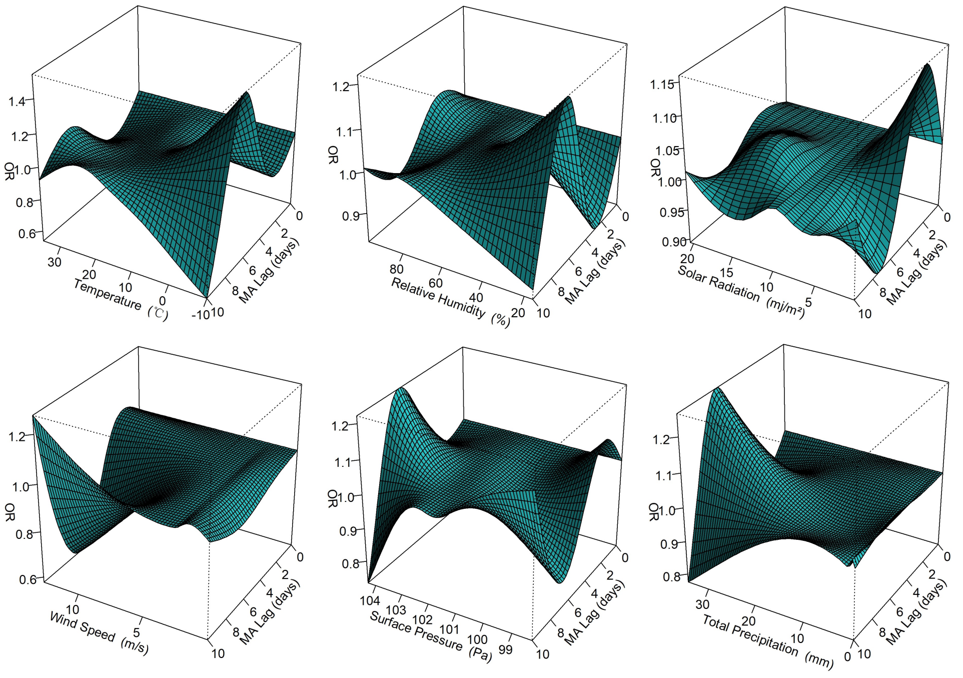


Figure S3: Three-dimensional plots for OR surface with ma-lag models and non-linear effect in DLNM.

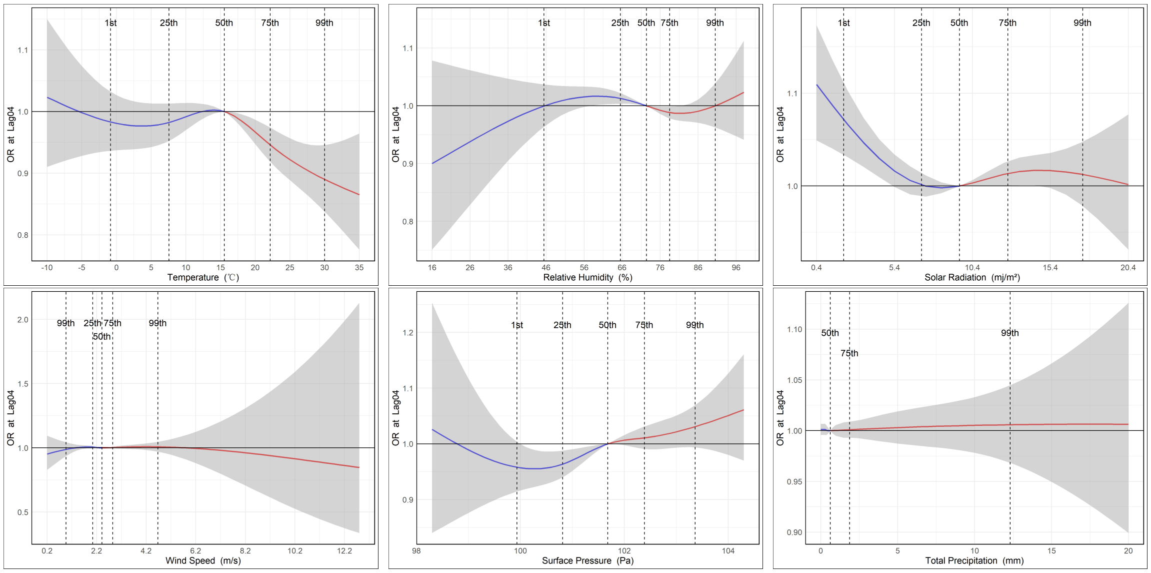


Figure S4: Exposure curves for non-linear relationship in the DLNM ma-lag04 model.

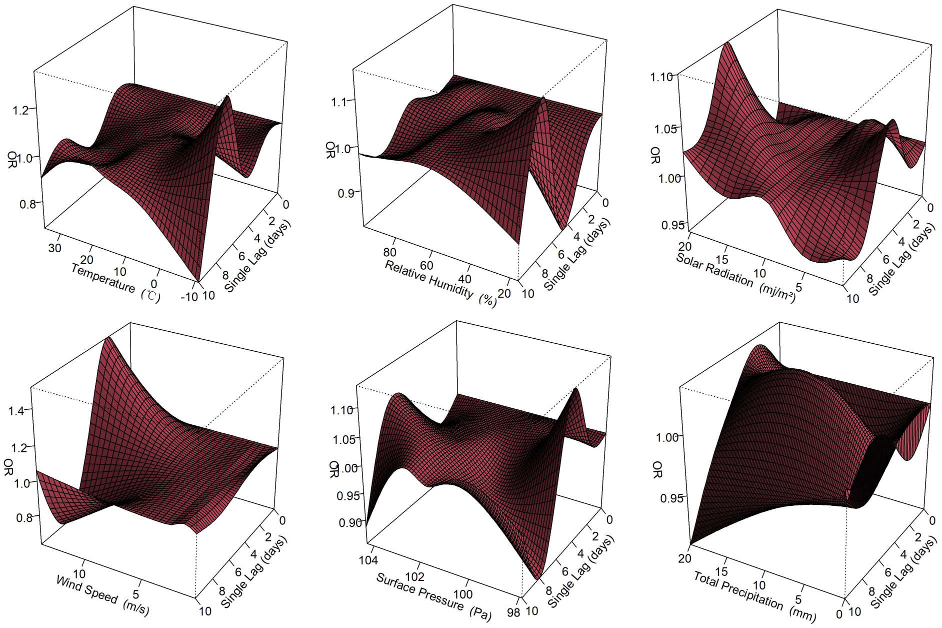


Figure S5: Sensitivity analysis with degrees of freedom 4. Three-dimensional plots for OR surface with single-lag models and non-linear effect in DLNM.

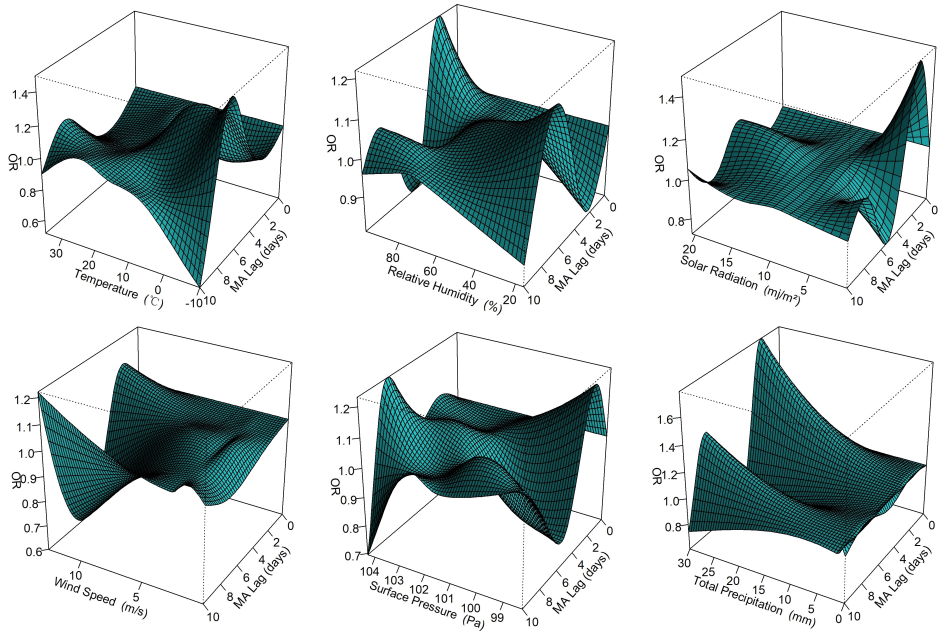


Figure S6: Sensitivity analysis with degrees of freedom 4. Three-dimensional plots for OR surface with ma-lag models and non-linear effect in DLNM.

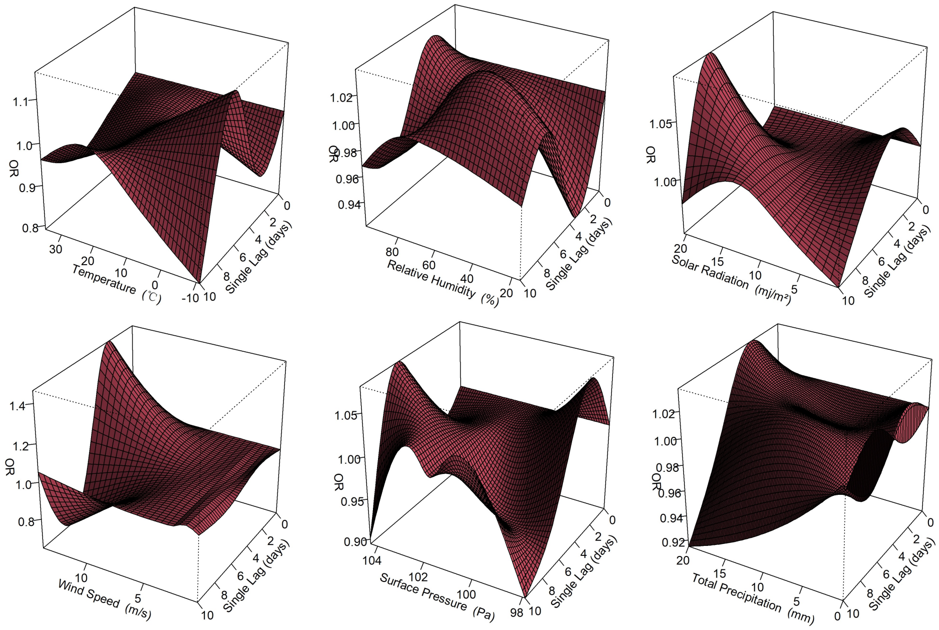


Figure S7: Sensitivity analysis with knots at 33rd and 66th percentile. Three-dimensional plots for OR surface with single-lag models and non-linear effect in DLNM.

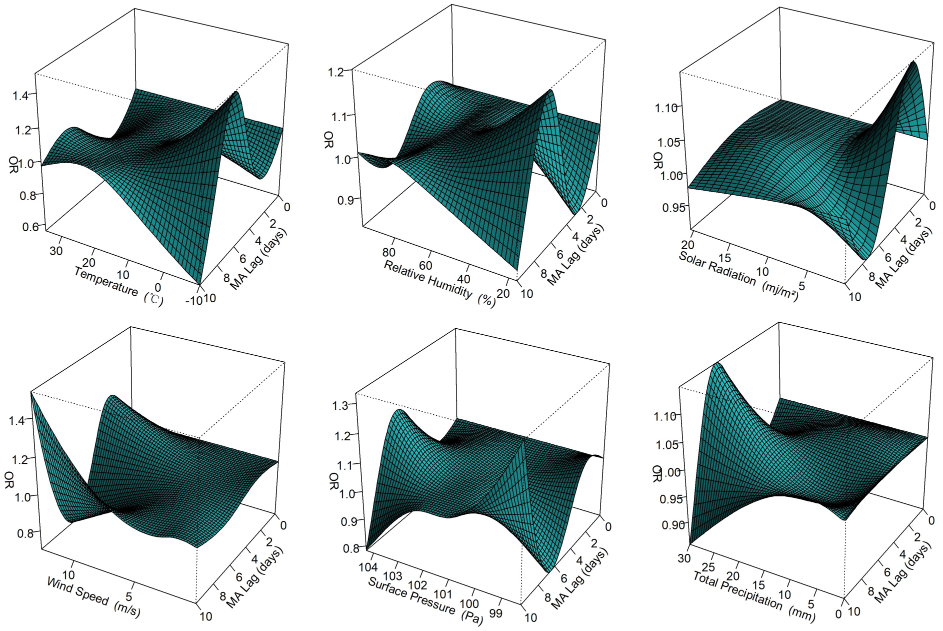


Figure S8: Sensitivity analysis with knots at 33rd and 66th percentile. Three-dimensional plots for OR surface with ma-lag models and non-linear effect in DLNM.

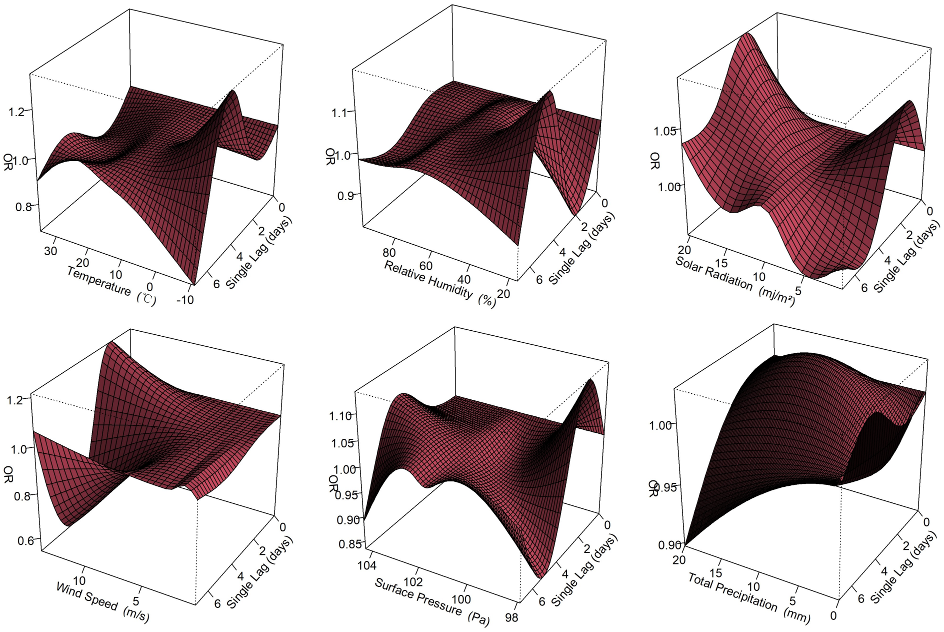


Figure S9: Sensitivity analysis with max lag days of 7. Three-dimensional plots for OR surface with single-lag models and non-linear effect in DLNM.

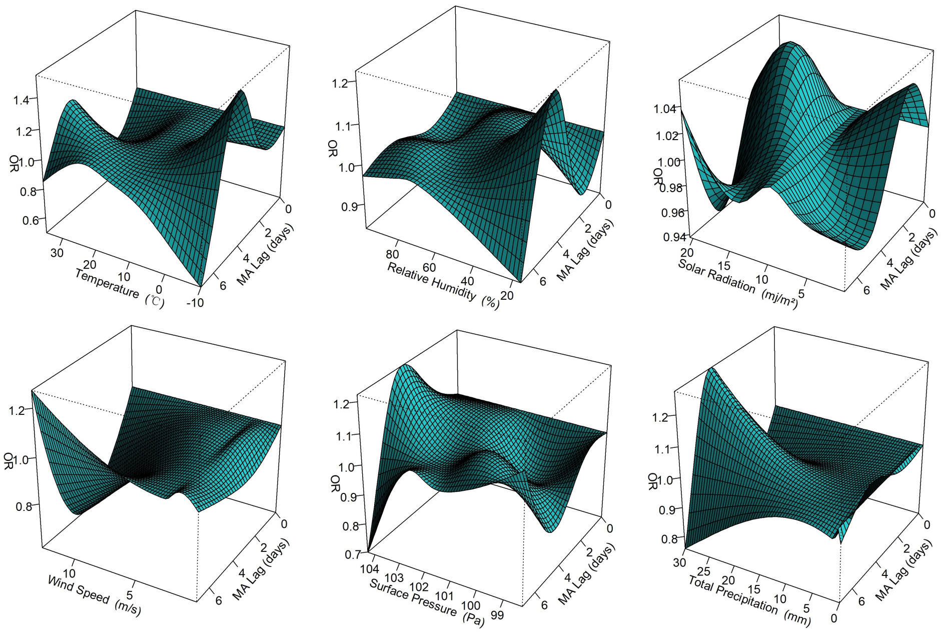


Figure S10: Sensitivity analysis with max lag days of 7. Three-dimensional plots for OR surface with ma-lag models and non-linear effect in DLNM.

### 2 Tables

Table S1: Characteristics of the study subjects in Jiangsu Province, China, from 2005 to 2023.

| Characteristic | | Number of cases (%) |
|---|---|---|
| Age | 0–2 | 555 (1.7) |
| | 3–9 | 28231 (84.6) |
| | 10–14 | 3791 (11.4) |
| | 15+ | 788 (2.4) |
| Season | Spring | 10952 (32.8) |
| | Summer | 6868 (20.6) |
| | Autumn | 6070 (18.2) |
| | Winter | 9475 (28.4) |
| Gender | Male | 20362 (61.0) |
| | Female | 13003 (38.9) |
| Time period | 2005–2010 | 4998 (15.0) |
| | 2011–2019 | 22498 (67.4) |
| | 2020–2023 | 5869 (17.6) |

Table S2: Summary of interaction single-lag3 models with OR, REOI, AP and their 95% CIs. The protective roles are modified with negative signs to be the risk roles, interpreted as the higher risk with lower values of exposures.

| Exposure pairs | 1st exposure's OR (95% CI) | 2nd exposure's OR (95% CI) | Interaction's OR (95% CI) | REOI (95% CI) | AP (95% CI) | S (95% CI) |
|---|---|---|---|---|---|---|
| (Temp, −RH) | 1.006 (0.997, 1.016) | 1.001 (0.999, 1.004) | 1.0002 (1.0001, 1.0003) | 0.0002 ($7\times10^{-5}$, 0.0004) | 0.0002 ($7\times10^{-5}$, 0.0004) | 1.0275 (1.0114, 1.0439) |
| (−Temp, −WS) | 1.012 (1.006, 1.017) | 1.021 (0.996, 1.047) | 1.0009 (0.9995, 1.0023) | 0.0011 (−0.0006, 0.0031) | 0.0012 (−0.0006, 0.0029) | 1.0357 (1.0079, 1.0642) |
| (−Temp, SRR) | 1.012 (1.006, 1.019) | 1.008 (0.999, 1.017) | 0.9998 (0.9994, 1.0003) | $-3\times10^{-5}$ (−0.0006, 0.0005) | $-3\times10^{-5}$ (−0.0005, 0.0005) | 0.9987 (0.9741, 1.0239) |
| (−Temp, SP) | 1.172 (0.913, 1.506) | 1.055 (1.004, 1.109) | 0.9984 (0.9960, 1.0009) | 0.0076 (−0.0020, 0.0172) | 0.0061 (−0.0006, 0.0128) | 1.0333 (1.0085, 1.0588) |
| (−Temp, −TP) | 1.009 (1.005, 1.014) | 1.012 (0.999, 1.026) | 1.0003 (0.9997, 1.0009) | 0.0004 (−0.0003, 0.0011) | 0.0004 (−0.0003, 0.0012) | 1.0194 (0.9970, 1.0423) |
| (−RH, WS) | 1.003 (1.001, 1.006) | 1.029 (0.963, 1.099) | 1.0006 (0.9997, 1.0015) | 0.0007 (− 0.0004, 0.0017) | 0.0007 (−0.0004, 0.0017) | 1.0214 (1.0109, 1.0319) |
| (−RH, SRR) | 1.004 (1.001, 1.007) | 1.006 (0.985, 1.026) | 1.0001 (0.9998, 1.0003) | $9\times10^{-5}$ (−0.0002, 0.0004) | $9\times10^{-5}$ (−0.0002, 0.0004) | 1.0099 (0.9949, 1.0252) |
| (−RH, −SP) | 1.120 (0.993, 1.264) | 1.024 (0.941, 1.116) | 1.0010 (0.9998, 1.0023) | 0.0042 (−0.0111, 0.0196) | 0.0037 (−0.0088, 0.0162) | 1.0292 (0.9693, 1.0927) |
| (−RH, −TP) | 1.005 (1.003, 1.006) | 1.005 (0.947, 1.066) | 1.0001 (0.9994, 1.0007) | $7\times10^{-5}$ (−0.0008, 0.0009) | $7\times10^{-5}$ (−0.0008, 0.0010) | 1.0075 (0.9603, 1.0570) |
| (−WS, SRR) | 1.022 (0.996, 1.049) | 1.004 (0.996, 1.011) | 0.9981 (0.9957, 1.0006) | −0.0018 (−0.0043, 0.0007) | −0.0018 (−0.0043, 0.0007) | 0.9306 (0.8688, 0.9967) |
| (WS, SP) | 1.600 (0.460, 2.567) | 1.103 (1.055, 1.153) | 0.9953 (0.9831, 1.0075) | 0.0545 (−0.1429, 0.2501) | 0.0305 (−0.0428, 0.1038) | 1.0761 (1.0154, 1.1405) |
| (−WS, −TP) | 1.003 (0.989, 1.016) | 1.003 (0.996, 1.009) | 0.9989 (0.9969, 1.0010) | −0.0010 (−0.0031, 0.0011) | −0.0010 (−0.0031, 0.0011) | 0.8333 (0.3434, 2.0219) |
| (SRR, SP) | 1.087 (0.738, 1.601) | 1.085 (1.034, 1.138) | 0.9992 (0.9954, 1.0031) | 0.0065 (−0.0281, 0.0412) | 0.0055 (−0.0215, 0.0326) | 1.0379 (0.9434, 1.1420) |
| (SRR, TP) | 1.008 (1.004, 1.012) | 1.001 (0.996, 1.007) | 0.9990 (0.9979, 1.0002) | −0.0010 (−0.0020, 0.0002) | −0.0010 (−0.0020, 0.0002) | 0.9058 (0.8098, 1.0133) |
| (SP, TP) | 1.082 (1.050, 1.114) | 1.361 (0.869, 2.133) | 0.9969 (0.9924, 1.0001) | 0.0250 (−0.0187, 0.0687) | 0.0170 (−0.0053, 0.0392) | 1.0564 (1.0291, 1.0844) |

Table S3: Summary of interaction ma-lag04 models with OR, REOI, AP and their 95% CIs. The protective roles are modified with negative signs to be the risk roles, interpreted as the higher risk with lower values of exposures.

| Exposure pairs | 1st exposure's OR (95% CI) | 2nd exposure's OR (95% CI) | Interaction's OR (95% CI) | REOI (95% CI) | AP (95% CI) | S (95% CI) |
|---|---|---|---|---|---|---|
| (Temp, −RH) | 1.011 (0.996, 1.025) | 1.003 (0.999, 1.006) | 1.0002 (1.0000, 1.0004) | 0.0003 ($5\times10^{-5}$, 0.0005) | 0.0003 ($5\times10^{-5}$, 0.0005) | 1.0198 (1.0128, 1.0269) |
| (−Temp, −WS) | 1.019 (1.011, 1.028) | 1.077 (1.027, 1.129) | 1.0039 (1.0013, 1.0064) | 0.0057 (0.0014, 0.0102) | 0.0053 (0.0015, 0.0090) | 1.0596 (1.0438, 1.0758) |
| (−Temp, SRR) | 1.008 (0.999, 1.017) | 1.025 (1.012, 1.039) | 0.9996 (0.9989, 1.0003) | 0.0006 ($-7\times10^{-5}$, 0.0012) | 0.0006 ($-6\times10^{-5}$, 0.0012) | 1.0169 (1.0010, 1.0331) |
| (−Temp, SP) | 1.283 (0.929, 1.772) | 1.090 (1.019, 1.165) | 0.9975 (0.9943, 1.0006) | 0.0220 (−0.0027, 0.0468) | 0.0158 (0.0017, 0.0298) | 1.0591 (1.0251, 1.0941) |
| (−Temp, −TP) | 1.009 (1.004, 1.015) | 1.028 (1.004, 1.053) | 1.0007 (0.9996, 1.0017) | 0.0009 (−0.0004, 0.0022) | 0.0009 (−0.0004, 0.0023) | 1.0253 (1.0057, 1.0453) |
| (−RH, WS) | 1.006 (1.000, 1.012) | 1.006 (0.856, 1.183) | 1.0002 (0.9980, 1.0024) | 0.0003 (−0.0029, 0.0035) | 0.0003 (−0.0029, 0.0035) | 1.0257 (0.9558, 1.1008) |
| (−RH, SRR) | 1.006 (1.001, 1.011) | 1.004 (0.972, 1.037) | 1.0000 (0.9996, 1.0004) | $5\times10^{-6}$ (−0.0006, 0.0006) | $5\times10^{-6}$ (−0.0006, 0.0006) | 1.0005 (0.9439, 1.0604) |
| (−RH, −SP) | 1.180 (0.974, 1.430) | 1.042 (0.911, 1.192) | 1.0015 (0.9997, 1.0034) | 0.0095 (−0.0279, 0.0471) | 0.0077 (−0.0201, 0.0357) | 1.0429 (0.9487, 1.1465) |
| (−RH, −TP) | 1.006 (1.004, 1.008) | 1.004 (0.942, 1.070) | 0.9999 (0.9992, 1.0007) | $2\times10^{-5}$ (−0.0011, 0.0011) | $2\times10^{-5}$ (−0.0011, 0.0012) | 1.0022 (0.9120, 1.1013) |
| (−WS, SRR) | 1.061 (0.997, 1.128) | 1.001 (0.984, 1.017) | 0.9939 (0.9881, 0.9997) | −0.0063 (−0.0137, 0.0009) | −0.0060 (−0.0127, 0.0007) | 0.8965 (0.8406, 0.9561) |
| (−WS, SP) | 1.008 (0.995, 1.014) | 1.107 (0.903, 1.069) | 0.9811 (0.9579, 0.9944) | 0.0218 (0.0367, 0.0067) | 0.0202 (0.0344, 0.0060) | 1.1678 (1.1046, 1.2346) |
| (WS, −TP) | 1.005 (0.978, 1.031) | 1.005 (0.987, 1.023) | 1.0032 (0.9967, 1.0096) | 0.0031 (−0.0032, 0.0097) | 0.0031 (−0.0032, 0.0097) | 1.3299 (0.5831, 3.0333) |
| (−SRR, SP) | 1.061 (0.583, 1.930) | 1.095 (1.023, 1.172) | 0.9992 (0.9933, 1.0051) | 0.0049 (−0.0444, 0.0543) | 0.0042 (−0.0359, 0.0444) | 1.0317 (0.8504, 1.2515) |
| (SRR, TP) | 1.016 (1.009, 1.023) | 1.005 (0.987, 1.023) | 0.9989 (0.9969, 1.0008) | −0.0011 (−0.0028, 0.0007) | −0.0010 (−0.0027, 0.0007) | 0.9502 (0.8986, 1.0048) |
| (SP, TP) | 1.103 (1.061, 1.147) | 1.749 (0.743, 4.121) | 0.9948 (0.9859, 1.0028) | 0.0668 (−0.0723,0.2060) | 0.0348 (−0.0087, 0.0783) | 1.0783 (1.0401, 1.1179) |

Table S4: Subgroup analysis of age group, gender, and periods, based on ma-lag 04 model.

| Groups | | Temperature OR (95% CI) | Relative humidity OR (95% CI) | Solar radiation OR (95% CI) | Wind speed OR (95% CI) | Surface pressure OR (95% CI) | Total precipitation OR (95% CI) |
|---|---|---|---|---|---|---|---|
| Age | 0–6 | 0.992 (0.982, 1.001) | 0.995 (0.991, 0.998) | 1.013 (1.002, 1.023) | 1.006 (0.966, 1.048) | 1.063 (0.993, 1.139) | 0.999 (0.987, 1.010) |
| | 6+ | 0.988 (0.980, 0.995) | 0.991 (0.988, 0.994) | 1.019 (1.010, 1.027) | 0.992 (0.960, 1.026) | 1.180 (1.118, 1.245) | 0.980 (0.970, 0.990) |
| Gender | Male | 0.986 (0.979, 0.993) | 0.993 (0.991, 0.996) | 1.013 (1.005, 1.022) | 1.014 (0.981, 1.048) | 1.146 (1.086, 1.210) | 0.989 (0.979, 0.998) |
| | Female | 0.994 (0.985, 1.003) | 0.991 (0.988, 0.995) | 1.020 (1.010, 1.031) | 0.971 (0.932, 1.013) | 1.110 (1.037, 1.188) | 0.987 (0.975, 0.998) |
| Time period | 2005–2010 | 0.992 (0.977, 1.008) | 0.992 (0.986, 0.998) | 1.009 (0.990, 1.028) | 1.035 (0.964, 1.111) | 1.098 (0.981, 1.229) | 0.997 (0.975, 1.020) |
| | 2011–2019 | 0.990 (0.983, 0.997) | 0.994 (0.992, 0.997) | 1.013 (1.005, 1.021) | 0.990 (0.959, 1.022) | 1.114 (1.057, 1.174) | 0.990 (0.981, 0.999) |
| | 2020–2023 | 0.985 (0.973, 0.998) | 0.985 (0.980, 0.990) | 1.037 (1.021, 1.054) | 0.971 (0.913, 1.032) | 1.235 (1.123, 1.358) | 0.974 (0.958, 0.991) |
